# Among patients with pneumonia-related sepsis, do those with mental health problems have different radiographic findings than those without mental health problems?

**DOI:** 10.64898/2026.04.30.26352126

**Authors:** Htet Ng, Daisy Puca, Jonathan Perla, Mark Richman

## Abstract

**Introduction:** Pneumonia is a common source of sepsis and carries significant morbidity and mortality. Prior research suggests patients with severe mental illness may receive disparate care and experience worse outcomes from infectious and cardiovascular conditions. Whether mental illness influences the radiographic presentation of pneumonia-related sepsis, which can guide antimicrobial selection and clinical decision making, remains poorly-understood.

**Methods:** This retrospective study examined chest imaging findings in 202 adult septic patients with respiratory source infection presenting to a large tertiary care emergency department between December 2017 and December 2019. Patients were stratified by the presence (n = 51, 25.2%) or absence (n = 151, 74.8%) of a comorbid mental illness, defined broadly to include neurocognitive, mood, personality, psychotic, and substance use disorders. Radiographic findings, including parenchymal abnormalities, pleural effusions, and laterality, were systematically abstracted from the medical record and compared between groups using Chi-squared testing.

**Results:** Patients with mental illness were more likely to have at least one radiographic finding (84% vs. 74%), though this difference did not reach statistical significance (p = 0.14). The average number of findings per patient was nearly identical between groups (3.14 vs. 3.12, p = 0.38), and no individual radiographic feature differed significantly between cohorts.

**Discussion:** These findings suggest that, contrary to our hypothesis, mental illness may not be associated with the radiographic appearance of pneumonia in septic patients. Larger, diagnosis-specific studies are needed to evaluate whether specific psychiatric subgroups exhibit distinct imaging patterns.

## Introduction

Pneumonia is a leading cause of morbidity and mortality worldwide, particularly when complicated by sepsis.

^1^ The radiographic appearance of pneumonia is often assessed using chest plain film radiography or computed tomography (CT) scans.^2^ These imaging modalities allow clinicians to visualize the anatomic extent of the infection and guide treatment decisions.

Our group is conducting a study comparing the quality of care of septic patients with vs. without mental illness, as determined by the percent of each group who meets the Surviving Sepsis Campaigns’ 3- and 6-hour bundles. Previous research has demonstrated patients with severe mental illness and cardiovascular disease or diabetes are 30% less likely to receive recommended cardiovascular or diabetes care, including hospitalization, diagnostic tests, appropriate medications, and invasive procedures.^3^ A national cross-sectional study in Sweden found patients with severe mental illness had an increased risk of death associated with influenza/pneumonia and sepsis.^4^

Antibiotic therapy for sepsis can be tailored according to the likely or confirmed organ involved, and which organisms are most-likely based on demographic, clinical, or radiographic findings. For example, a finding of pneumonia accompanied by lung cavitation increases the likelihood of methicillin-resistant staphylococcus aureus (MRSA) compared with pneumonia with lobar infiltrates only.^5^ Likewise, right lower lobe infiltrates are more-likely to be from aspiration pneumonia, because the right main stem bronchus is wider and straighter than the left main stem bronchus.^6^

Recent studies indicate the radiographic appearance of pneumonia varies among populations, including those with vs. without mental illness.^7,8^ This current study aims to add to this nascent literature by further examining whether pneumonia-related sepsis has different radiographic findings in patients with vs. without mental illness. We hypothesize individuals with pre-existing mental illness who develop sepsis-related pneumonia may exhibit distinctive radiographic patterns compared to those without mental illness. Alterations in mental health and associated behavior (eg, aspiration owing to dementia)^9^ may impact the presentation and progression of pneumonia in septic patients, thereby influencing the radiographic appearance.

## Materials and Methods

Northwell Health is a 22-hospital health system largely operating in Long Island and New York City. Long Island Jewish Medical Center (LIJMC) is a 583-bed tertiary care teaching hospital serving a racially and socio-economically diverse population. The adult ED sees approximately 100,000 patients per year. Members of LIJ’s Department of Quality Services collect and submit sepsis data to state and federal agencies. Patients with a contraindication to bundle components (pregnant or those with advanced directives for non-aggressive medical care) are removed from the sepsis database.

Patients were included in this study if they attended the LIJ adult ED between December 2017 and December 2019 and were in the LIJ Department of Quality Service’s sepsis database. Patients with sepsis and any of the ICD-10 codes in Appendix A for mental health conditions in the broad categories of neurocognitive disorders (eg, dementia), mood disorders (eg, depression), personality disorders (eg, borderline), psychotic disorders (eg, schizophrenia), and substance use disorders (eg, alcohol) were included.

We filtered all 584 patients in the database to determine which had sepsis owing to pneumonia, as indicated by the field “Site of infection” having the value “Respiratory.” We then separated those patients by those with vs. without mental illness, as indicated by the patient having any of the ICD-10 codes in Appendix A. We did a chart review to determine what radiographic study(ies) the patient had and categorized the appearance of any findings, as follows:

1. Side
2. Site (eg, which lobe)
3. Parenchymal findings:
  a. Abscess
  b. Airspace opacification
  c. Air bronchogram
  d. Air-fluid level
  e. Aspiration
  f. Atelectasis
  g. Cavitation
  h. Consolidation (includes pulmonary edema)
  i. Ground-glass opacity
  j. Interstitial pattern
  k. Lobar or segmental involvement
  l. Nodules
  m. Pleural effusion
  n. Pleural thickening
  o. Pleuritis
  p. Silhouette sign

Chi-squared testing was used to compare the proportion of patients with vs. without mental illness who demonstrated each radiographic finding. Statistical significance was set a priori at p < 0.05.

This project was deemed to fall under the realm of performance improvement by the Northwell Health Institutional Review Board (IRB #: 24-0076), and, therefore, did not require formal ethics oversight.

## Results

A total of 202 septic patients with pneumonia-related sepsis were included in the analysis, of whom 51 (25.2%) had a comorbid mental illness and 151 (74.8%) did not. At least one radiographic finding was present in 84% (43 or 51) patients with mental illness and 74% (112 of 151) patients without mental illness. The between-group difference of 10.1% was not statistically-significant (95% CI: -3.7 to 20.8; p = 0.14). Among patients with at least one finding, the mean number of findings per patient was nearly identical between groups (3.14 vs. 3.12; p = 0.38). No statistically-significant differences were identified between groups in any individual radiographic finding category (**Table 1**). As illustrated in Figure 1, parenchymal-only findings were the most common category in both groups, followed by effusions alone, combined parenchymal and effusion findings, and no findings, with the distribution remaining broadly similar across both cohorts. Regarding laterality, bilateral involvement was the most frequently observed pattern in both groups, followed by left-sided and right-sided involvement, with no statistically-significant differences between groups across any laterality category (**Figure 2**).

**Table 1.**
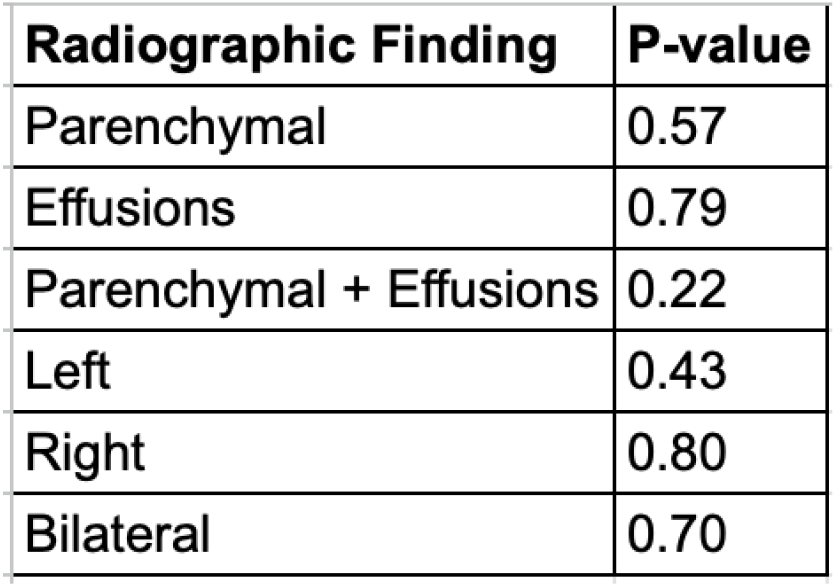
Chi-Squared Analysis of Radiographic Findings Between Septic Pneumonia Patients With and Without Mental Illness.

**Figure 1.**
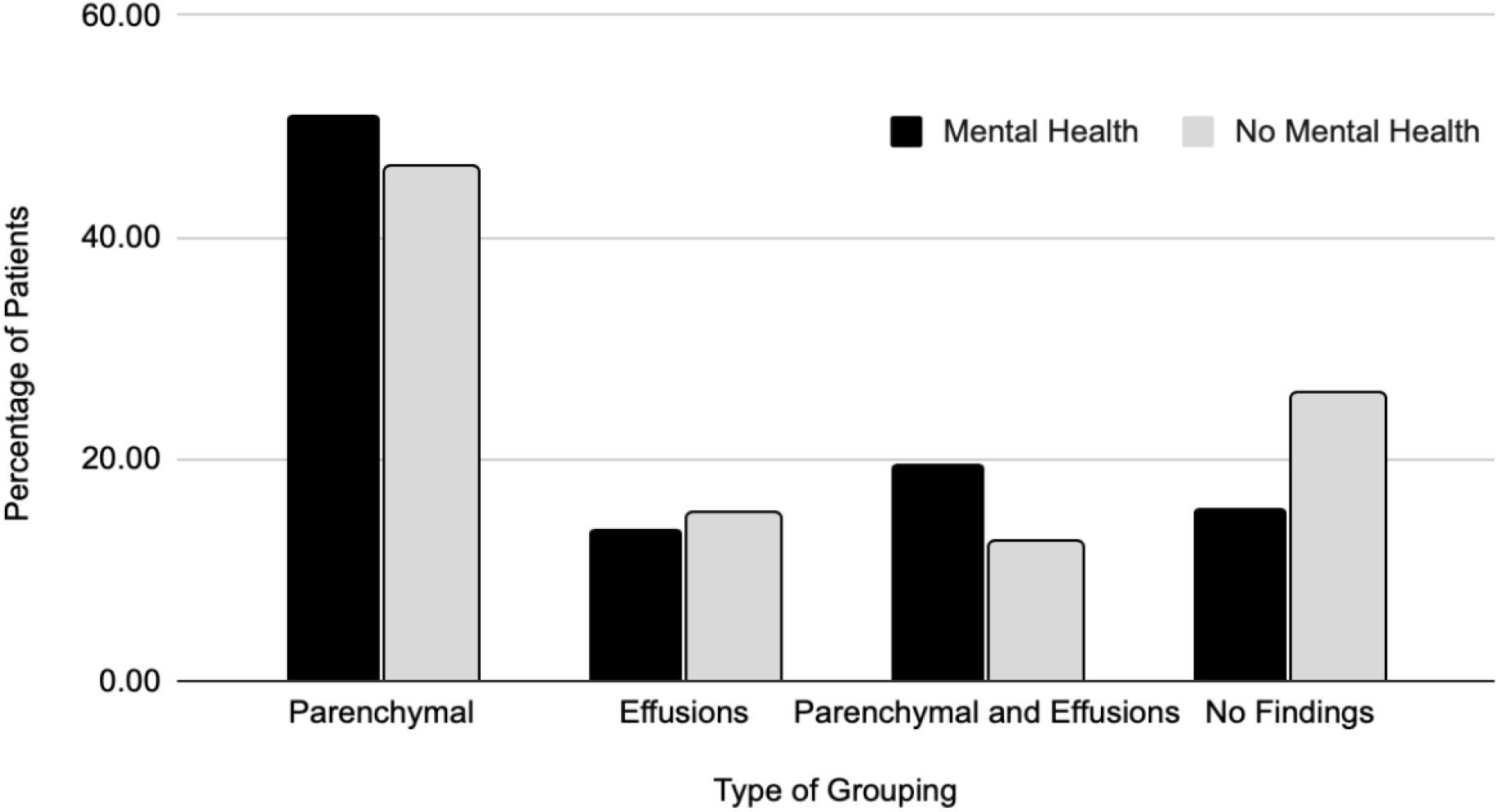
Distribution of Radiographic Finding Categories Among Septic Patients with Pneumonia, Stratified by Mental Illness Status

**Figure 2.**
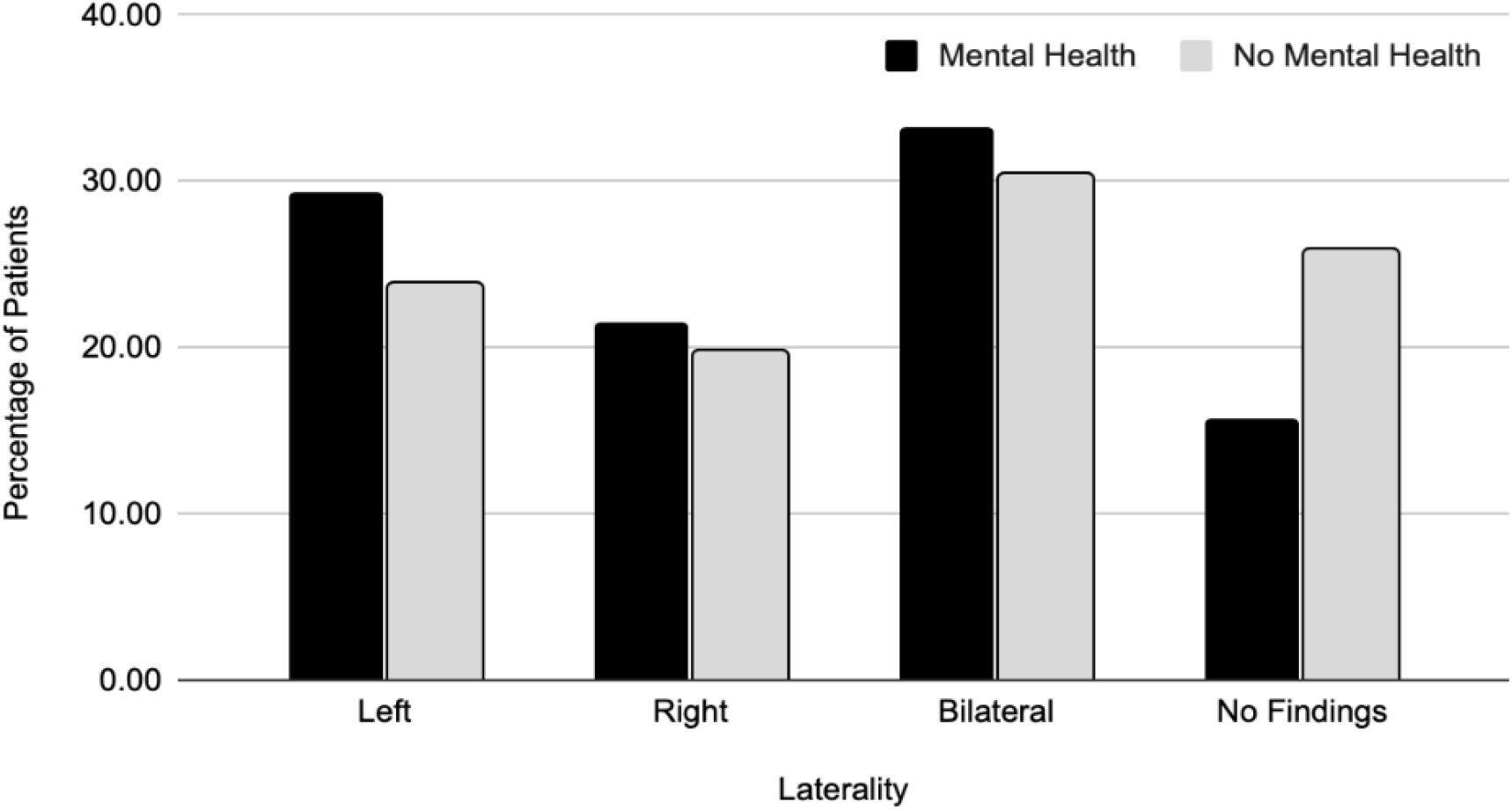
Laterality of Pneumonic Infiltrates Among Septic Patients with Pneumonia, Stratified by Mental Illness Status

## Discussion

The aim of this study was to determine if patients with pneumonia-related sepsis show unique radiographic patterns dependent on the presence of concomitant mental disorder. In contrast to our hypothesis, our analysis did not find statistically-significant differences between the two cohorts. Our data shows that patients with and without mental illness had statistically-similar radiographic findings, including parenchymal alterations, pleural effusions, and the laterality of the infection (left, right, or bilateral). These results counter previous research, such as the Chen et al. study,^8^ which hypothesized that pleural effusions and multi-lobar involvement may be more common in people with schizophrenia.

### Limitations

The most-significant limitation of this study is the relatively-small sample size (n = 202). The study may not have had enough power to identify radiographic differences because there were only 51 individuals in the mental illness cohort. Also, the study’s wide definition of “mental illness,” which includes cognition, mood, personality, and psychotic disorders, may have obscured certain radiographic abnormalities associated with a specific subcategory. For example, behaviors that are explicitly linked to dementia may demonstrate different imaging than behaviors linked to drug use disorders; such findings might require a larger sample size to identify.

## Data Availability

All data produced in the present study are available upon reasonable request to the authors

## Appendix A ICD-10 codes for severe mental illness

ICD-10 codes for severe mental illness
| Neurocognitive Disorders |
| --- |
| Alzheimer's (G30.0) |
| Alzheimer's late onset (G30.1) |
| **Alzheimer's specified NEC (G30.8)** |
| **Alzheimer's with behavioral disturbance (G30.9)** |
| **Delirium (F05)** |
| **Dementia (F03.90)** |
| **Dementia with aggressive behavior (F03.91)** |
| **Frontal Dementia (G31.09)** |
| **Hepatolenticular degeneration (<u>E83.01</u>)** |
| **HIV infection (F02 and B20)** |
| **Huntington's disease (F02 and G10)** |
| **Lewy bodies (G31.83)** |
| **Multiple etiologies (F03)** |
| **Neurosyphilis (A52.17 )** |
| **Parkinson's with behavioral disturbance (F02 and G20)** |
| **Senile with acute confusional state (F05)** |
| **Wilson's disease (E83. 01)** |
| **Vascular (acute onset) (mixed) (multi-infarct) (subcortical) (F01.50)** |
| **Vascular (acute onset) (mixed) (multi-infarct) (subcortical) with behavioral disturbance (F01.51)** |
| **Vitamin B12 deficiency (D51.9)** |
| **Mood Disorders** |
| **Bipolar disorder (F31.\*)** |
| **Major depression (F32.0)** |
| **Personality Disorders** |
| **Antisocial personality disorder (F60.2)** |
| **Borderline personality disorder (F60.3)** |
| **Psychotic Disorders** |
| **Catatonia (F06.1)** |
| **Delusional disorder (F22)** |
| **Schizoaffective disorder, bipolar type (F25.\*)** |
| **Schizophrenia, paranoid (F20.0)** |
| **Schizophrenia, disorganized (F20.1)** |
| **Substance Use Disorders** |
| **Alcoholic (F10.97)** |
| **Alcoholic with dependence (F10.27)** |
| **Alcohol withdrawal (F10.239)** |
| **Cocaine use, unspecified with unspecified cocaine-induced disorder (F10240)** |
| **Inhalants with dependence (F18.27)** |
| **Psychoactive drug (F19.97)** |
| **Sedative/hypnotic/anxiolytic dependence w withdrawal, unsp (F13.239)** |
| **Sedatives, hypnotics or anxiolytics (F13.97)** |
| **Sedatives, hypnotics or anxiolytics with dependence (F13.27)** |
| **Volatile solvents (F18.97)** |

